# On the heterogeneity of infections, containment measures and the preliminary forecast of COVID-19 epidemic

**DOI:** 10.1101/2020.05.12.20096792

**Authors:** Ichiro Nakamoto, Weiqing Zhuang, Sheng Wang, Yan Guo

**Author notes:** Corresponding author: Ichiro Nakamoto Author.

## Abstract

**Background:** As of May 1, 2020, there had been over three million of officially confirmed cases of novel coronavirus (COVID-19) infections reported worldwide. The pandemic originated from a severe acute respiratory syndrome coronavirus 2 (SARS-CoV-2), a virus similar to severe acute respiratory syndrome (SARS). The dynamics of the pathogen incurred the incidence of the unidentified cases that were potentially substantial in magnitude. Unparalleled extensive measures, either in terms of medical quarantine or non-medical containment, were taken to deplete the growth of infected population and thereafter settle down the outbreak. We aimed to estimate the gap in sizes and peak dates between the confirmed and unconfirmed, and how containment measures impacted the dynamic trajectory of the COVID-19 in Japan. We performed simulations and desired to provide meaningful insight for the upcoming responses to the outbreak, for which much still remained to be unknown.

**Methods:** To examine the differentiation between identified and unidentified cases, and how heterogeneity of medical quarantine and non-medical control were associated with the advancement of the outbreak dynamics, we employed the susceptible-infected-removed-contained (SIR-C) model that derived from the basic SIR concept where the target population was divided into three differentiated compartments: (S)usceptibles (be subject to infections), (I)nfected (confirmed infections) and (R)emoved (not in the procedure of transmission due to the reason of either recovered or died). By applying the transmission model to the latest outbreak data in Japan, we established the least-squares fitted estimates parameters and the epidemiological trajectory of the COVID-19 pandemic. In compliance with the estimated framework, we simulated the ongoing trend of outbreak in Japan by calibrating the potential changes in measures ceteris paribus starting April 7, the date when the state of emergency was declared. We assumed a variety of settings and simulated how the heterogeneity in containment shifted the subsequent advancement of the outbreak.

**Findings:** The epidemiologically estimated outcomes with least-squares fitting indicated a gap between the confirmed and unconfirmed cases in terms of size and peak dates. The saturation size of the reported infections was comparative to the unidentified infections in magnitude contingent on the duration of infection (DOI). However, peak dates for the former delayed by nearly two-and-half months. The declaration of state of emergency incurred changed patterns in social behaviors and sub-exponential growth of the outbreak. Medical and non-medical measures were effective in controlling the outbreak of COVID-19. By assuming a changed pattern of containment measures since April 7, a diminishing growth of infections and reduced saturation of cases were to be observed, accompanied by an earlier arrival of peak dates. However, the modelled effects of quarantine and control measures vary with the unclear infectiousness and the attributes of containment.

**Interpretation:** As the number of infected cases, especially of those asymptomatic, mild symptomatic and infection-route-unknown cases, was growing over time, it was of importance to verify the assumption of the potential existence of the gap in size between those already identified and those not. Our analysis reinforced this by quantifying this magnitude. The trend curve of reported cases differentiated from unidentified cases with a time-lagged effect. Containment measures, if followed effectually, would probably help reduce the spread of epidemic. Our simulations suggested that 1% of growth in rate of non-medical containment could approximately predate the peak by 18 days and reduce the peak size by one-thirds. Commensurate level of containment effect was to be reached when the rate of medical quarantine was increased ten-folds. We thus projected that changes in interventions could lead to an earlier peak date but reduced peak size, which could be flattened by calibrating the interventions gradually. Limitations of our research include the uncertainties in the estimates of duration of infection and the reproduction number.

**Funding:** Philosophy and Social Sciences Association in FuJian, Science and Technology Development Center of the Ministry of Education.

**Research in context:** *Evidence before this research:* COVID-19 pandemic was reported to emerge in late 2019 and cases were reported ever since. China adopted multiple options of efforts including medical quarantine and non-medical measures, such as isolation of cases, physical distancing, contact tracing, school closure and workplace shutdown, at a national level to break down the spread of COVID-19. By significantly decreasing the likelihood of close proximity person-to-person contact, these measures took effect and by the end of March 2020 the reported cases lowered dramatically. Japan identified its first case of COVID-19 on Jan 22, 2020 and dynamic exponential-like growth of cases was observed since. The investigation of epidemiological feature for containment measures in other countries attracted much attention; in contrast, the trajectory for Japan remained to be unclear. We searched web of science and science direct for research published in English from 2019 up to May 6, 2020, with the terms “ coronavirus control or containment measures” in combination with “Japan” and identified 6 and 5 results respectively. Relatively little was known about the gap in size and in peak time between the confirmed and unconfirmed in Japan; how containment measures influenced its trajectory of COVID-19 since the state of emergency was declared; and how the shift of the pandemic was influenced by the containment.

*Added value of this study:* The measures studied were divided into two broad ranges in Japan: medical quarantine and non-medical containment. Medical quarantine typically corresponded to cases with severe symptoms that need urgent care; in comparison, non-medical measures consisted of a wider variety of control by complementing the effect of quarantine and corresponded to cases such as asymptomatic, pre-symptomatic, mild symptomatic patients and the susceptibles. We found that the declaration of state of emergency incurred social behaviors change and was in connection with subsequent sub-exponential growth of cases. Our study implied the essential existence of gap, subsequently quantified its asymptotic size and its peak timing between the identified and the unidentified. Time-delaying effect in saturation was meaningful in that it provided the information to understand the intrinsic intra-correlation of an epidemic like COVID-19. The outcomes of simulation implied that early containment measures which linked to a reduced and predated peak, were crucial to control further spread. A dynamic transmission model was employed to assess the potential trajectory starting the point of emergency declaration.

*Implications of all the available evidence:* The efficacy of containment has been identified in China and induced changes of behaviors in individuals responsive to the pandemic where asymptomatic, mildly symptomatic and infection-route-unknown cases were not to be ascertained. This might be of great importance for developing control strategies for currently yet to be convergent communities or possibly subsequent cyclical outbreaks of COVID-19. It was of difficulty to estimate the accurate gap in practice; however, asymptotic measures could deliver information that tended to be ignored. Medical quarantine and non-medical measures were at work during the COVID-19 outbreak and the test of heterogeneity on its quantitative and qualitative efficacy in different countries might assist to uncover more on the unknown of this pathogen. Parameter-fitted simulations could provide decision makers with scenarios not reflected in the available information and thus the insights on timely and prioritized decisions even in the uncertainties such as COVID-19.

## Introduction

The COVID-19 epidemic incurred viral respiratory diseases and pneumonia outbreaks worldwide. Countries such as France found that it was already spreading in December 2019, a month before the official first cases in the country. It was projected that COVID-19 epidemic might have started much earlier than assumed.^1^ As of May 1, 2020, confirmed infections reached 3,145,407, including 221,823 deaths (i.e., average mortality rate of nearly 7·05%) based on the report by WHO.^2^ The accurate number of unconfirmed cases were to be ascertained. This has potentially changed the past concept of local and sporadic outbreaks of epidemics to an extensive and compounding cycle of response and recovery.^3^ A variety of unrivalled medical quarantine and non-medical containment measures has been employed in response to the SARS-CoV-2 pandemic.^4^ While comprehensive adverse effects on the infected and biological attributes of the pathogen were to be uncovered, countries and communities with early isolating of cases, curtailing person-to-person contacts, physical distancing, tracing close contact in combination with hygiene practices (e.g., utilization of masks or disinfectant) have obtained meaningful control and the practice may be of interest to other communities where divergent growth of the infections was still under way and where later growth tended to drastically shift from earlier baseline at a pace out of expectation. ^4^,^5^

The dynamics of human-to-human transmission risk correlated with the external interventions coupled with interactions of individuals and other time-contingent factors that might impact the trajectory.^6^ In the unpredicted occurrence where information on the transmission fluctuation and routes was incomplete, prompt containment, timely screening and isolation evolved to be crucial in control of the spread risk of the pandemic. Medical (i.e., pharmaceutical) quarantine was used in numerous affected countries, and non-medical (i.e., non-pharmaceutical) measures such as physical distancing and contact tracing effectively influenced the trajectory of progression in shrinking the outbreak of COVID-19. Local residents were typically called upon to stay home when able and to perform physical distancing and hygiene practice when not. Control measures aimed at reducing the contact in the population delayed the peak and reduced the ultimate size of the epidemic. ^7,8^

Discrete spreading events in connection to a later explosion of spread had been identified for past SARS outbreak and were not exceptional for COVID-19 epidemic as well, unmanageable and random mass movement deteriorated the spread. Hence, early shutdown of the transmission routes of these discrete events was of importance to diminish the further spread.^9,10^ Another concern was that subclinical transmission typically caused by asymptomatic or pre-symptomatic individuals might worsen the pandemic. WHO estimated that COVID-19 outbreak was still at its preliminary stage, thus it was vital to gain updated understanding of the potential effects of available control measures in terms of medical and non-medical perspective on the evolution trajectory of the pandemic.

If infectious individuals were to be identified timely and effectively then the efficacy of measures could be increased significantly and thus concentrating effort on the quarantining of the most infectious cases would be at work than mere random control.^9^ However, such kind of mechanism was compromised during a pandemic in which transmission established prior to the onset of symptoms.^10^ As the rate of spread hinged partially on the dynamics of the pathogen such as basic reproduction number Ro and the duration of infection, which were defined as the average secondary infections for each index case and the inverse of removal rate respectively.^11^ And the greater of these values, the more difficult for the outbreak to settle down. Hence, stringent implementation of measures was critical in effective interruption of the chains of transmission.^12^

Outbreaks of SARS characterized marked differences between the affected regions in total infections and epidemic duration, even for those where outbreaks started and identical control measures were enacted simultaneously.^13^ Knowledge was still insufficient about COVID-19. Preventing further spread and controlling subsequent occurrence of pandemic remained to be a global priority.^14^ And the existence of asymptomatic and pre-symptomatic infections complicated the trajectory of epidemic.^15^ Human-to-human transmission was confirmed in other places of Japan since its first identified case.^16^ Countries could migrate to be worldwide epicenters of outbreak unless substantial health interventions at a variety of levels were implemented instantly. Large scale of asymptomatic and pre-symptomatic transmission in the absence of public health interventions would induce international seeding and subsequent local establishment of epidemics inevitable.^17,18^

To quantify the gap between the confirmed and unconfirmed cases in terms of sizes and peak dates and anticipate how the trend of COVID-2019 spread was influenced by the containment, we first performed least-squares fitted parameters analysis based on outbreak surveillance data in Japan and then forwarded to the simulations of the spread trajectories contingent on the fitting outcome by assuming containment changes triggered since the emergency declaration.

## State of emergency over COVID-19

A state of emergency declared by the government as of April 7, 2020 for Tokyo, Osaka and five other prefectures to curb the outbreak of COVID-19 effective through May 6, 2020 after an alarming rise of infections was observed and extended to nationwide later on. A cut of at least 70 percent in human-to-human contact was expected. By May 4, it was further extended until the end of that month. The declaration enabled prefectures to employ stronger preventive measures, ranging from instructing citizens to stay home, avoid “3Cs” (i.e., closed spaces where crowds gathered in close proximity), restricting the operation of schools and other facilities. It hinged greatly on self-compliance of individuals and no legal penalties for noncompliance up to that point. Government issued a series of economic assistance by then, including a uniform subsidy of 10 thousand JPY to each individual and support programs for firms in response to the epidemic. ^19^

## Methods

### SIR-C model

The dynamics of the transmission of COVID-19 over time are to be governed by the set of differential equations as follows:

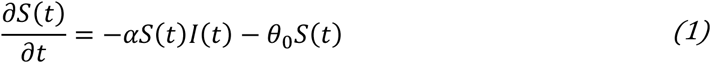

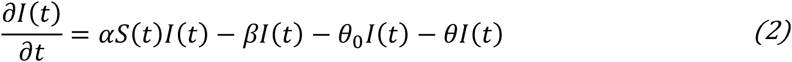

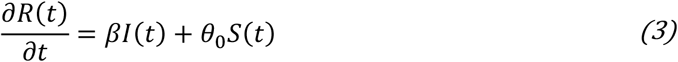

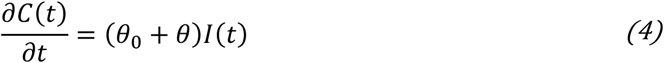

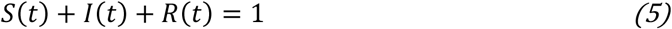

This SIR-C model derives from the standard SIR. *S*(*t*)*, I*(*t*) *and R*(*t*) denote the time-contingent fraction of the susceptibles, infected and removed cases respectively. The total number of population is N with initial infection of one individual and initial removed individual of zero. The size of N was acquired from the computation by Statistics Bureau of Japan as of April 1, 2020 and fixed to 125·96 million by assumption. The outbreak data used the official surveillance information from the Ministry of Health, Labor and Welfare of Japan. To account for the potential effects on the population resulting from the containment, one new time-contingent compartment *C*(*t*) was introduced to quantify the dynamic fraction being medically quarantined (e.g., severe cases) and non-medically contained (e.g., susceptibles, asymptomatic or mild cases). The sum of both containment is termed as the social health effort in outbreak control. In general, *I*(*t*) is the fraction of infections in population unable to be exactly identified in practice accounting for aforementioned factors. The assumption is that *C*(*t*) maintains a positive and close relation with the empirically identified naive cases and thus approximates to the reported cases. *θ*_0_ denotes the rate of non-medical control such as staying at home, physical distancing, and hence in general captures the uniform effect on the population including asymptomatic, mild-symptom individuals, susceptibles and removed individuals; in contrast, *θ* typically captures the effect of rate of medical quarantine on the infected with severe symptoms. Or equivalently, measures influence the susceptibles, asymptomatic, mild- and severe-symptom individuals in an identical way. Parameters *α* and *β* quantify the rate of transmission and removal respectively. For this set of first-order differential system, symbolic solution for eq. (1) is:

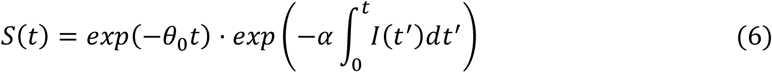

At its transmission stage, we assume that containment measures impact the progression of outbreak in a more dominant way such that the influence resulting from the transmission process is to be negligible. Consequently approximated solution for the susceptible simplifies to the following:^20^

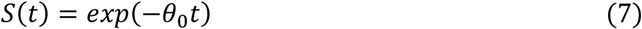

Substitute (7) into eq. (2) and derive the general solution for the practically unidentifiable fraction of infected as:

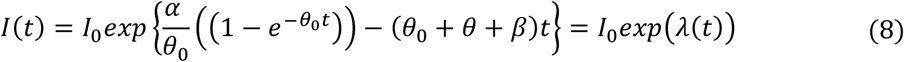

for the case *θ*_0_ ≠ *0* and:

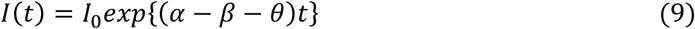

for the case where *θ*_0_ ≠ *0*, which catches the effect in which non-medical control exerts trivial or no effect at all, or target population is not to respond to the containment in a noticeable way. The trend of infected *I*(*t*) hinges on the parameters of transmission rate, removal rate, rate of non-medical or medical containment and the time. In the non-zero scenario, if 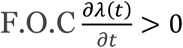, then unidentified infected *I*(*t*) increases over time. In contrast, if 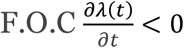 the opposite trend is to be observed. At the locus where 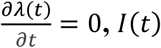 reaches its saturation (i.e., peak), from which we derive the relationship below:

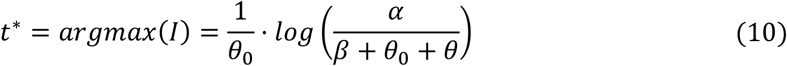

Where 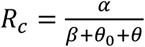 denotes the effective reproduction number when external control measures are to be enacted and 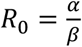 is the basic reproduction number of no containment respectively. Thus, when the rate of non-medical containment increases ceteris paribus, *R_c_* decreases accordingly. Similar trend applies to the case of medical quarantine ceteris paribus. Both types of reproduction number capture the subsequent infections for each index case on average prior to the removal from the transmission. Reduced reproduction number was associated with the slowed-down or ceasing spread of epidemic. When the rate of non-medical control measure *θ*_0_ is infinitely close to zero, the value of *t^*^* potentially approximates to a large value such that the spread is hard to diminish. In contrast, a larger value of *θ*_0_ implies greater effort in containment, leading to smaller value of *t^*^* and thus an earlier arrival of peak dates. Similar analysis applies to medical quarantine rate *θ*. To establish the fitted parameters, we employed the non-linear least-squares of Levenberg–Marquardt.^20^ Then a Taylor-series expansion around the point *t*:

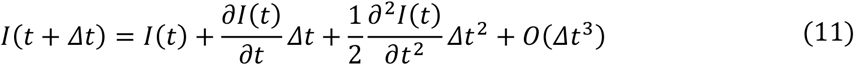

Where *Δt* is set as the step-size for the asymptotic computation. There are two types of intrinsic errors involved in this approach: First, the truncation of the Taylor series incurs error that limits the ultimate accuracy of the model. Second, utilization of the approximation of *I*(*t*) given by the previous iteration when computing *I*(*t + Δt*) generates an additional disturbance that may accumulate over successive iterations, and hence affects the quantitative sensibility of the method. The approximate number of unconfirmed infections at time *t* can be determined by:

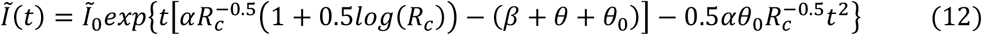

And the inherent association between time-adjacent unidentified infections is given by:

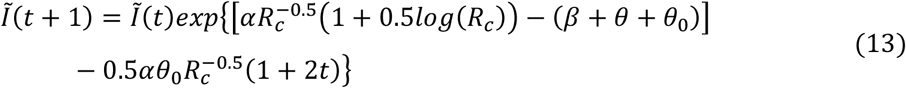

In contrast, confirmed cases are to be asymptotically decided by the relation below: ^20^

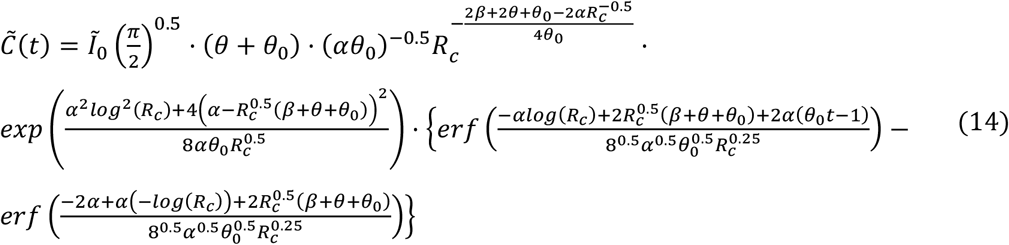

To estimate and simulate, part of the parameters was attained from published literature as presented in Table, for the remaining we least-squares fitted in calibration to outbreak data of Japan. The estimates utilized the cases from January 22 to May 3. The effect inherent in the containment was to be reflected by the fitting outcome, and analysis was performed to evaluate the fitting between discretely observed infections and the fitted curves. In the second scenario, since state of emergence was formally declared as of April 7, we assumed changed patterns in medical quarantine and non-medical containment respectively starting that day and simulated the trajectory of outbreak beyond the point contingent on the least-squares fitted parameters ceteris paribus. Research estimated that the mean basic reproduction number of SARS-CoV-2 was to range between 2 and 7.^11,22^ The practice in China had also shown that a value of 6.2 for the basic reproduction number qualitatively worked well.^20^ The effective reproduction number was estimated to be 2.48 in our study. Transmission dynamics of COVID-19 caused it difficult to identify and target risk groups. The virus was highly infectious and had a long but still uncertain transmission window as partial infections were ascertained to start prior to the onset of symptoms. According to announcement by WHO, the incubation period of COVID-19 ranged from 1 to 14 days with median estimates of 5 days. The removal rate consisted of the sum of recovery and mortality rate, it was known that variation existed in the removal procedure for the infected contingent on the status of sickness and the potential lasting health problems resulting from COVID-19. The duration of infection was defined as the inverse of the removal rate, and heterogeneity was found in its distribution. The variation of duration of infection was estimated to be in a fuzzy range of [3d, 20d].^20^ However, this duration could potentially also vary in a narrower interval of [5d, 20d].^21^ We found that a 16-day of duration fitted the outbreak well. To check, we explored how the differentiation in duration of infection affected the trajectory of outbreak by increasing its values from 8 to 18 days with a step of 2-day increment. The least-squares estimate for *θ* was close to the boundary, which implied that the medical quarantine was not at work in its equilibrium and a low capacity of healthcare was to be assumed.

|  | **Value** | **References** |
| --- | --- | --- |
| Base reproduction number without any containment: $R_0 = \frac{\alpha}{\beta}$ | 6.2 | Maier et al<sup>20</sup> |
| Transmission rate: $\alpha$ [d<sup>-1</sup>] | 0.39 | Calculated from $R_0$ and $\beta$ |
| Duration of infection: $\frac{1}{\beta}$ [d] | 16 | Assumed |
| Medical quarantine rate: $\theta$ | 7.6e-10 | Least-squares Fitted |
| Non-medical containment rate: $\theta_0$ | 0.016 | Least-squares Fitted |
| Effective reproduction number with containment: $R_c = \frac{\alpha}{\beta + \theta + \theta_0}$ | 2.48 | Least-squares Fitted |
| **Table: Parameters of the SIR-C model** |  |  |

The typical scenarios in which containment is to be employed in response to an epidemic can be summarized in their simplest (Figure 1). This applies to a setting in which the capacity or quality of healthcare is not at full performance or the capacity is of relative tininess. In the case in which equilibrium of healthcare is not to be presumed, treatment of infections is to be differentiated depending on infectiousness, symptoms and the accommodation capacity of the healthcare system. Privilege is given to severely symptomatic cases and thus immediate medical quarantine is needed. In contrast, asymptomatic or mild-symptom patients are typically recommended to comply with the strategy of non-medical containment by staying home or at locally assigned places. The dynamics of COVID-19 updates this by either deteriorating to severe infections or recovering from it. Severe symptomatic patients might recover or die from the intense medical quarantine by chance. For the general population, non-medical containment is enacted such as the necessity of hygiene practice or physical distancing.

**Figure 1:**
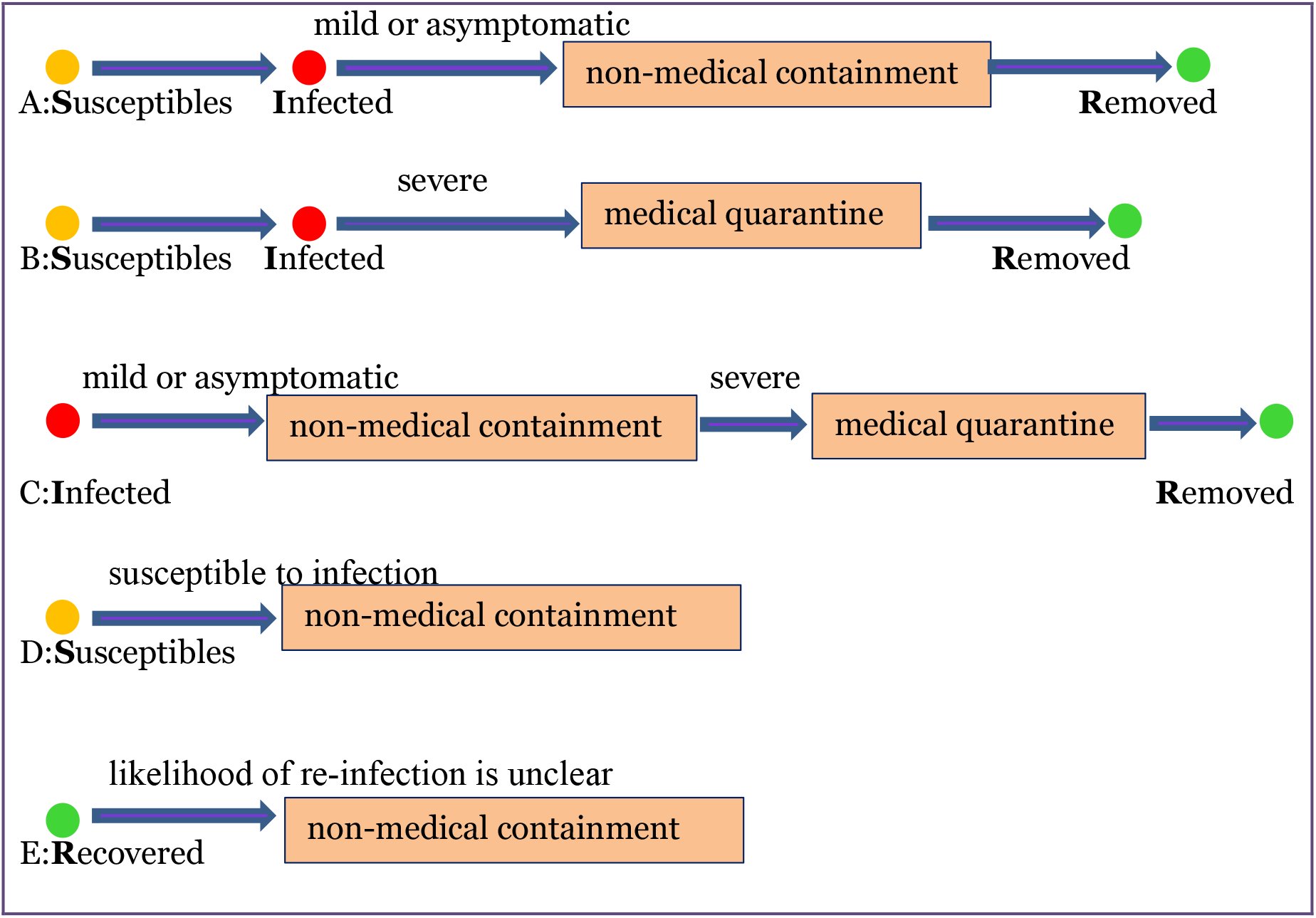
SIR-C model and procedure of containment contingent on symptoms, infectiousness and capacity of healthcare.

We divided the population into Susceptibles, Infected, Removed (either recovered or died) and Contained (either medically quarantined or non-medically contained) individuals. SIR-C= susceptible-infected-removed-contained. Each denotes one scenario that potential control is to be employed in response to the outbreak.

## Results

In the first scenario, we estimated the least-squares fitted results for the confirmed and unconfirmed cases with presumed effective reproduction number as well as duration of infection (presented in table) by using outbreak data of Japan from late January to early May (figure 2). In the subsequent scenarios, we then inherited all the fitted parameters but the rate of control with the utilization of an increment of one-day step. We calibrated the changes with respective to each containment starting the state of emergence to forecast the preliminary evolution trajectory of the outbreak beyond that point (figure 3; figure 4).

**Figure 2:**
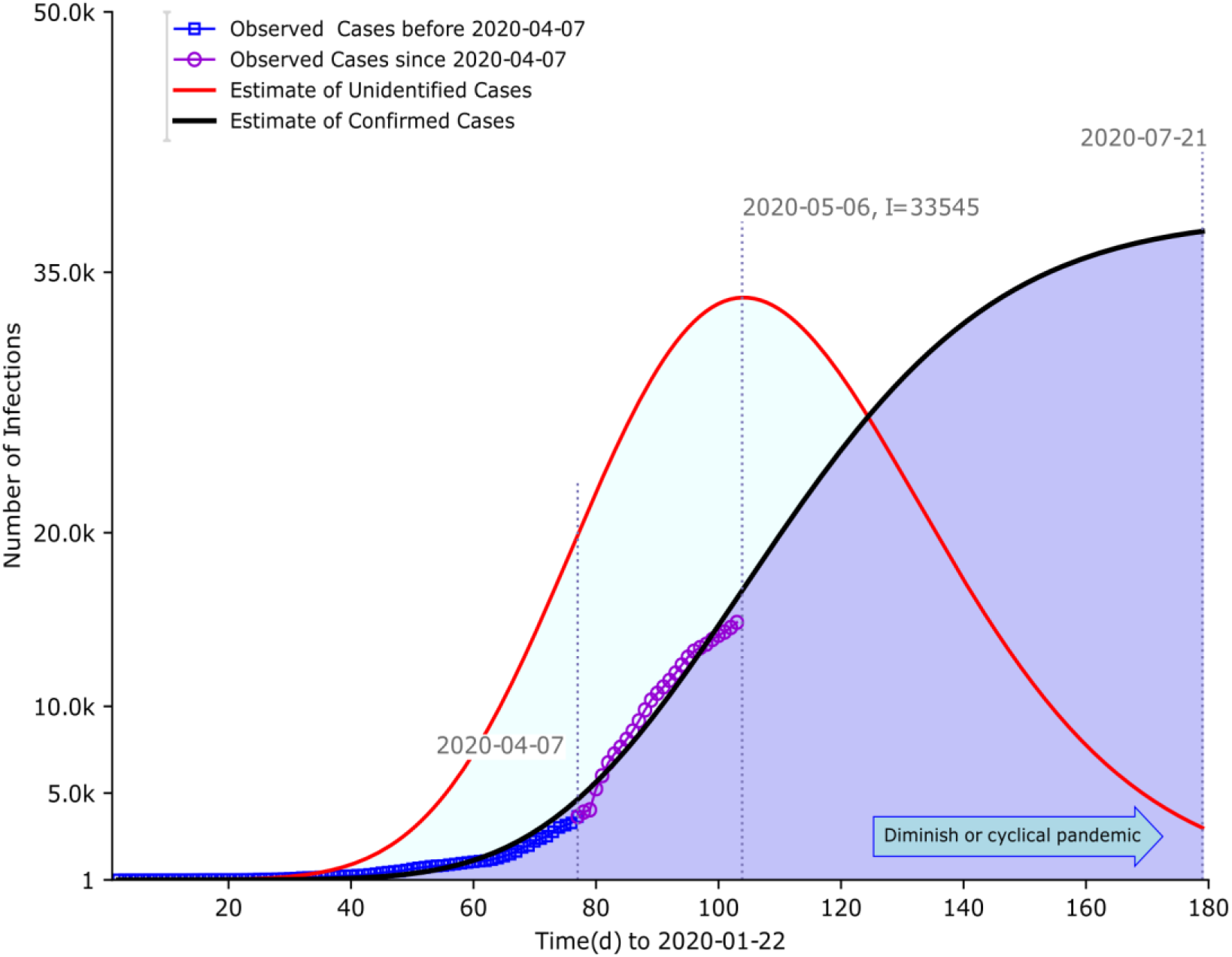
Estimates of confirmed *C(t)* and unconfirmed *I(t)* cases of COVID-19 in Japan from late January to early May. Parameters estimates were performed in calibration to reported cases predating May 4 based on least-squares fitting. Basic reproduction number without containment and duration of infection were set to be 16 days respectively. The effective reproduction number with containment was estimated to be 2·48. April 7 was the first date of state of emergence. The model predicted the inherent gap in size between the confirmed and the unconfirmed cases. The former peaked around July 21. In contrast, the latter saturated around May 6, almost two-and-half months earlier. Peak magnitude for both was at the commensurate level, approximated to 34000 cases. Lines of markers corresponded to the outbreak surveillance data of Japan from January 22 to May 3.

**Figure 3:**
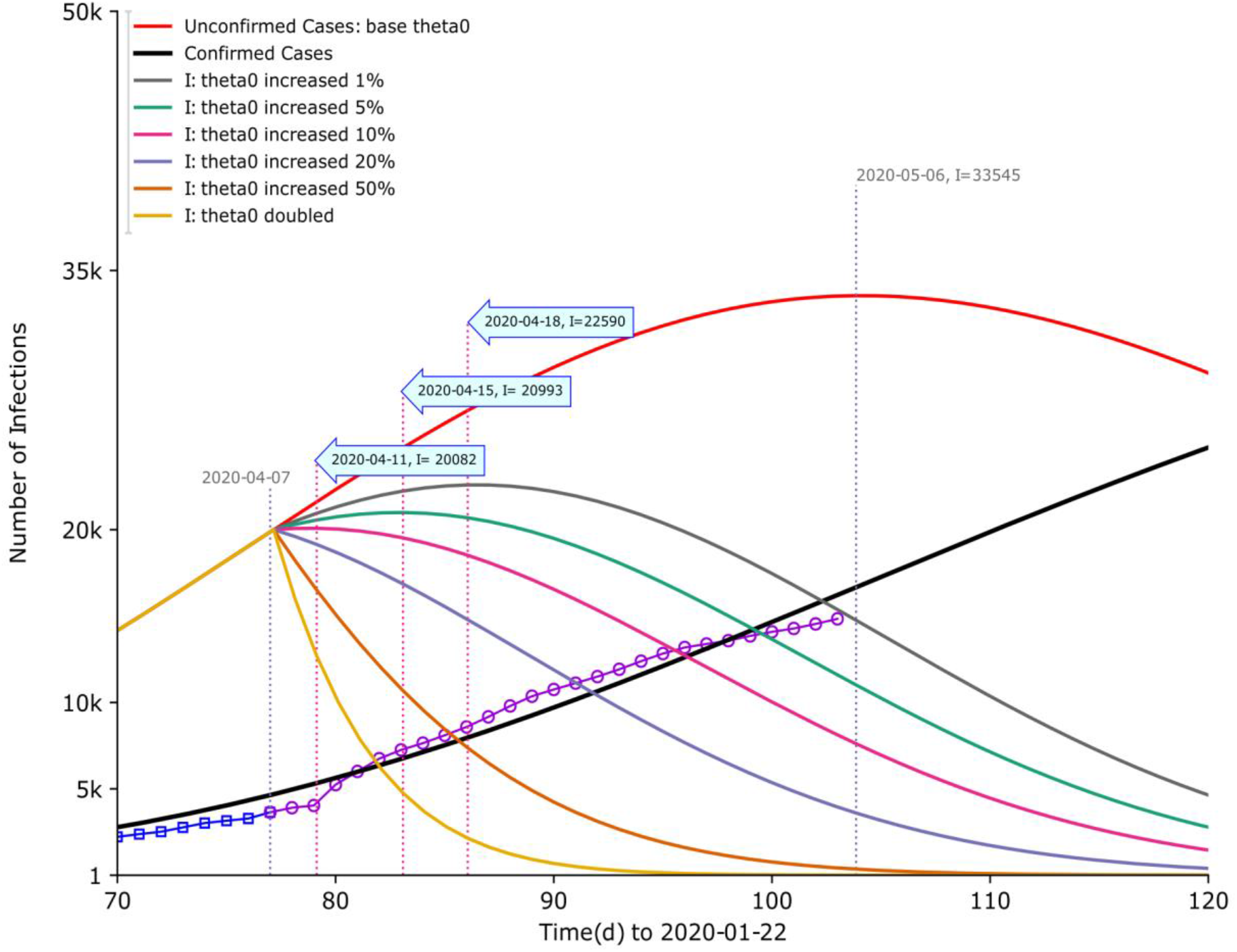
Simulations for confirmed and unconfirmed cases of COVID-19 with a change in rate of non-medical containment.

**Figure 4:**
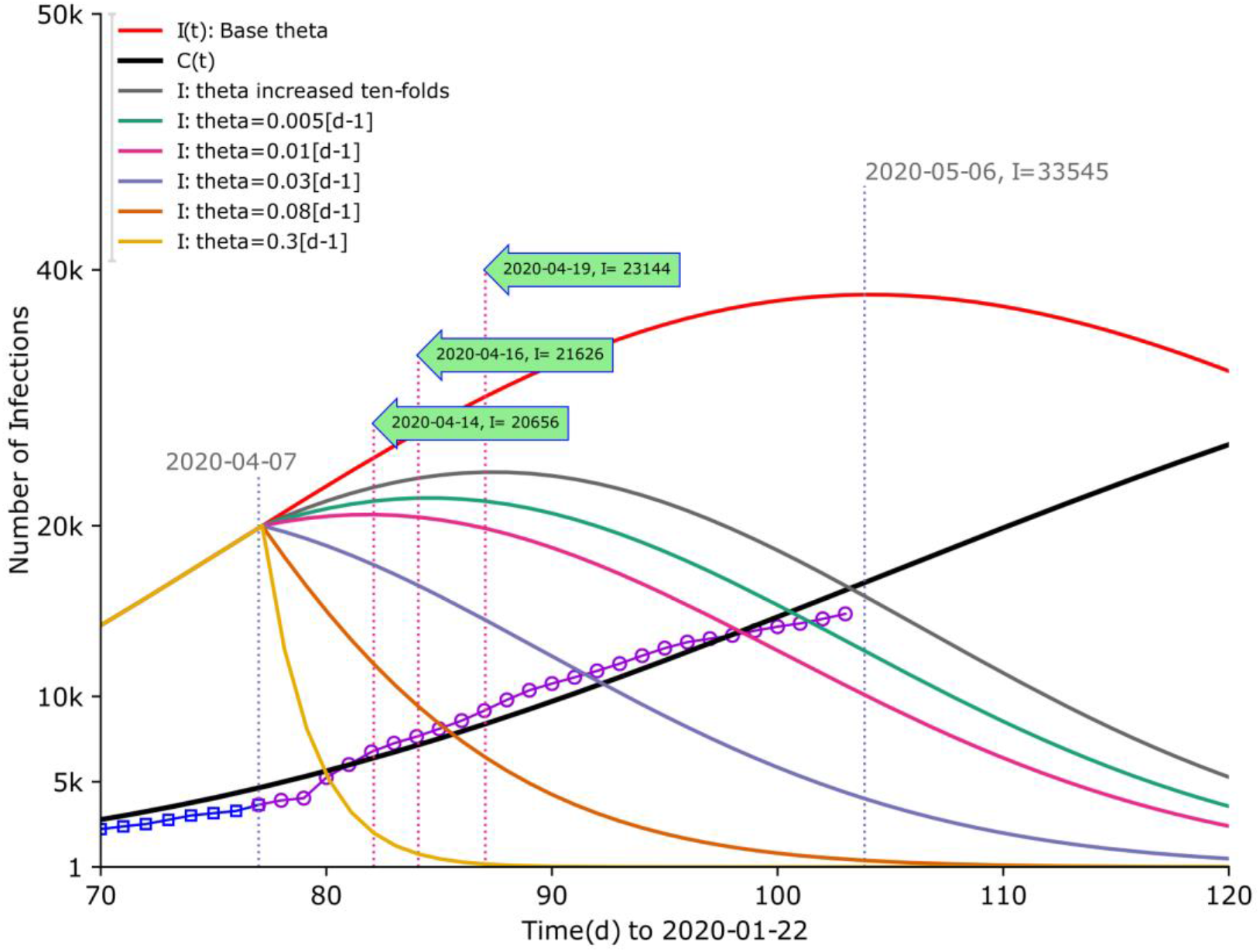
Simulations for confirmed and unconfirmed cases of COVID-19 with a change in rate of medical quarantine. All parameters in the simulations were fixed to the least-squared estimated values but the rate of medical quarantine. We assumed a sole change of quarantine rate occurred as of April 7. The data inside the colored arrows stood for the dates of maximum unrevealed infections and the magnitude. When rate of quarantine increased from 0·005 to 0·01 per day, dates of saturation occurred roughly three days earlier and reduction of almost 1500 cases. A decaying effect was to be observed at a point in between rate of 0·01 and 0·03. And a rate of 0·3 per day caused the outbreak to be controlled with efficacy by the end of three months since the first case was detected.

**Figure 5:**
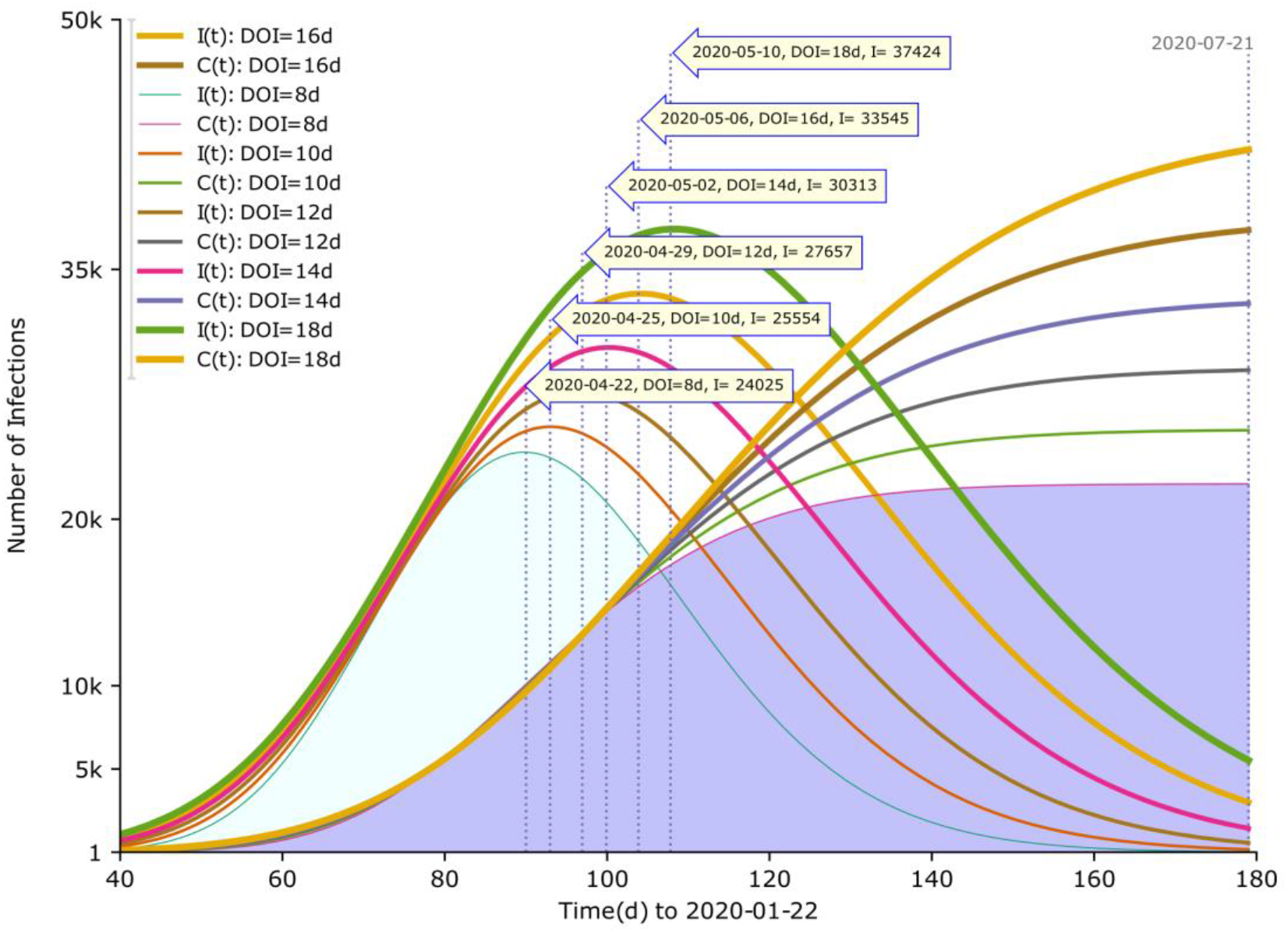
Effect of different duration of infection (DOI) on the confirmed and unconfirmed COVID-19 cases. Change DOI from 8 to 18 days by a two-day increment of step, the dates of saturation switched from April 22 to May 10 and the peak size increased from around 24 thousand to around 38 thousand cases. By late July, all confirmed infections tended to saturate at commensurate levels.

The estimates implied that the peaks of the unconfirmed and the confirmed did not coincide. The dates of peak for the former occurred around May 6 whereas it was delayed almost two-and-half months for the latter, which reached its peak around late July. In contrast, the ultimate saturation in magnitude for both was at a commensurate level, indicating approximately 34 thousand cases. At a time in between the two spikes, identified cases were to continue growing but unidentified cases started to decrease. Prior to the saturation of the unidentified, the size of gap was increasing over time but the opposite was to be observed after its summit was past and thereafter the pandemic faded gradually out or became the next starting point of recurrent outbreaks, as would be corrected by the dynamics of COVID-19. The estimate indicated that at a certain point after the state of emergence, the confirmed infections shifted to a sub-exponential or algebraic growth. Hence, it was projected that the state of emergence took effect and slowed down the initially exponential-like trajectory of outbreak (figure 2).

The simulations presented the association between the trend of unrevealed cases and the change in rate of non-medical containment ceteris paribus. Suppose an existence of change in containment rate starting April 7, in the case in which the rate of containment increased by 1%, the saturation date was projected to antedate nearly 18 days compared to no concurrent change at all, shifting to an earlier date of April 18, accompanied by a reduced peak size of 22590 cases (almost one-thirds decrease). It was to observe a greater downsize trend if the rate of containment increased by 5%, a near 37·4% of reduction in peak and date of saturation occurred almost three weeks earlier. The slowing-down effect was to shift to a decaying trend when rate of containment inflated by 20% or more, causing a faster pace of dying out of the outbreak. To achieve significant decrease in the unidentified cases such as 90% or more cut down in size by May 6, more stringent containment had to be enacted. It was postulated that increasing the rate of non-medical containment by 50% or more might potentially achieve this objective (figure 3).

All parameters in the simulations were time in-variant and inherited from the least-squared fitted values except the rate of non-medical containment. We assumed rate changes took place as of April 7, when the state of emergence was declared. The data inside the arrows denoted the dates of saturation and its magnitude.

In the scenario where rate of medical quarantine changed, the reduction in effective reproduction number would cause the pandemic to slow down or extinguish. This hinged on factors such as the capacity and quality of healthcare. Simulations conditional on this assumption illustrated the potential reduction in size as of April 7. If the rate of medical quarantine grew by 10-folds, it was projected that the peak date would come roughly 17 days earlier with a reduced peak size of 23144 infections. This is at a commensurate degree of cutting compared with one percent of growth in the rate of non-medical containment. The more increase in the rate, the greater curtailing effect on the peak size and earlier arrival of peak dates. A quarantine rate of 0·03 per day or larger could cause a decaying trend in the number of unidentified cases and thus a speedier extinguishment of the pandemic (figure 4). The curves also indicated that to achieve an at least 90% of depletion in cases, the rate of medical quarantine was to change at a value of 0·08 per day or more. Note that initial least-squares estimate of it was to fluctuate around the boundary, therefore this suggested a remarkable change in the quarantine.

To evaluate how differentiation in DOI impacted the trajectories of confirmed and unconfirmed cases, we altered its size from 8 to 18 days following a step of two-day increment. The difference of 10 days in DOI incurred 13399 (or around 56%) cases of increase for the unconfirmed, and the dates of peak relocated from April 22 to May 10, from which 18 days of shift was estimated. Similar trends were to be observed for the confirmed cases. By late July, all the curves of confirmed cases approximated to their saturation in magnitude asymptotically. Hence, the longer days of DOI, the longer of time to get to the peak of outbreak and the larger of peak size. This applied to trajectories of the confirmed and the unconfirmed identically. By asymptotical computation, every two-day increase of DOI was to linearly postpone the peak dates by three to four days; however, the growth in peak size did not present a similar linear trend. Initial growth of 1529 cases then rose to 2103 on the subsequent step, thereafter 2656, 3232 and 3879 respectively. Thus, in a similar vein when the DOI was to be lengthy, the saturation magnitude of the outbreak was projected to be sizeable. In contrast, when the DOI was relatively short, the spread was to be contained more rapidly. At low levels of DOI, the peak magnitude of the unconfirmed potentially surpassed the magnitude of the confirmed, and this trend shrank with the increase in DOI. Uncertainty in the dynamics of pathogen might partially contribute to this, however, the size of both generally maintained at comparable levels.

## Discussions

We found the quantitative and qualitative correlation between the revealed and unrevealed cases in peak dates and sizes contingent on presently available surveillance data. The dates of saturation for the unidentified did not occur simultaneously with the formally identified, implying the fundamental existence of the gap. It predated by nearly two-and-half months. In contrast, the peak size of both was at a comparative level. Both estimates hinged on the duration of infection. Prior to the saturation of the unidentified, the gap in size increased over time; in contrast, the opposite was to be observed when the summit was reached. A lengthier duration postponed the peak dates and enlarged the peak size accordingly, causing the outbreak more difficult and thus costly to settle down. The changes in the rate of medical quarantine and non-medical containment could quell the trend of unidentified cases and thus the spread. In a setting where the capacity of healthcare was presumed with relative stability, non-medical containment such as staying home or at local places, physical distancing and tracing close contact was to implement with priority. Our simulations implied that it could potentially incur a comparative level of effect when effectively followed. We projected that under an interval [8d,18d] for duration of infection, the peak size of COVID-19 outbreak in Japan would be of a range 24 to 38 thousand of unidentified infections, the peak dates of which would potentially fluctuate between late April and mid of May. Although a commensurate level of peak size could be expected for the confirmed cases, the time of saturation presented a delaying effect and converged until late May to late July hinging on the strength of the infectiousness. The declaration of state of emergency changed the initial exponential growth to the extent where peak date of outbreak would occur at a point around July. For the unidentified, this saturation might come at an earlier time, probably within the window of the emergency or by the end of May.

The trajectory of epidemic reflected the interactions of external containment strategies and the transmission factors coupled with changed behaviors in response to the outbreak. The heterogeneity in confirmed and unconfirmed cases, rate of quarantine and containment did influence the trajectory of outbreak. And this heterogeneity prospectively played its role during the first couple of months. One percentage increase in the rate of non-medical antedated the peak date by nearly 18 days and a size reduction of almost one-thirds. If the rate of medical quarantine increased by 10-folds, the peak date would arrive roughly 17 days earlier with an almost 33 percent cutting in peak size. We projected that confirmed cases were to saturate in late July or that time around and unidentified cases were to peak prior to that time, almost two-and-half months earlier. More stringent containment was of necessity in order to acquire greater diminishing effect on the outbreak.

The forecasts could potentially be impacted by other omitted factors such as capacity (e.g., pathogen testing capacity) and quality of healthcare, changing biological effects, social (e.g., the degree of shared sense of crisis in population) and spatial heterogeneities.^23,24^ Spatial variations such as structure of population mixing were found to exist, as the prefectures close to Tokyo were most populated in Japan. And for these communities, more stringent quarantine adherence was needed to a more flattened curve of outbreak.^25^ In the case resistance to drastic disease-control measures was at work, rising infection rates and mortality, coupled with scientific uncertainty about COVID-19, the curve of infections would fluctuate with larger uncertainty. We estimated the scenarios where medical quarantine and non-medical containment impacted the trends of COVID-19 pandemic. Preventing further transmission by decreasing the potential channels of infections and effective reproduction number. However, the feasibility of these strategies was to be compromised if the number of infected reached a threshold of the total population. One critical factor was how asymptomatic, pre-symptomatic and mild-symptomatic individuals responded to COVID-19 in regard to the feasibility of prevention of a second-time outbreak. Other factors that might impact the trajectory of the COVID-19 pandemic were not accounted for in our model as well. In the case where presumed assumptions were relaxed, the likelihood in connection with differentiated scenarios would increase. The simulation based on the fitted parameters indicated that the unidentified cases were generally larger than the reported cases. We investigated a range of scenarios where the heterogeneity of containment changed the trajectory of the transmission. As uncertainty existed in the factors such as the interval of infectiousness for asymptomatic, non-medical containment would be of importance to enhance the effect of control in combination with medical quarantine. The model could be modified to incorporate other unaccounted factors impacting the dynamics of the transmission, which would implicitly alter the trend of the COVID-19 outbreak. To avoid and control spread in randomness, the first couple of months might be of great importance. ^7,8^ As clinical knowledge of this novel pathogen and its dynamics accrues, it is feasible that outcomes will improve. It therefore will be of concern to revise these estimates as epidemics unfold.^26^

And because of the asymptotic approach utilized, qualitative evidence was to be established. The inherent relationship between the confirmed and unknown cases was asymptotically identified and hence its precision was up to the point where the inherent approach applied. The analysis could not differentiate the efficacy of specific containment, nor could it differentiate the effect for the asymptomatic, pre-symptomatic, symptomatic and infection-route-unknown compartment respectively. However, it reinforced other findings by showing that medical and non-medical control, when timely and successfully implemented, were feasible in decelerating or even diminishing the spread of the pandemic: with an earlier arrival of peak dates and reduced peak sizes. This could be informative when it was too lengthy for the outbreak to converge and when it was to accurately interpret the outbreak as “plus *α*” resulting from the dynamics. The meaning of this study partially lied in that we asymptotically identified the quantitative size of this. Containment measures were preferred to be performed at an earlier stage and the effectiveness has been identified in other countries and past outbreaks. Changes of behaviors were observed in response to the epidemic where asymptomatic, mild- and pre-symptomatic infections were not to be ascertained. The might be of great importance for developing control measures for presently widely spread secondary or future recurrent outbreaks of COVID-19.

While Japan may not be one of the countries with the highest infections or highest mortality rate of COVID-19 per capita worldwide to date, it has been listed as one of the chains transmissions resulting in extensive spread.^2^ Multiple important lessons emerged in that integration of healthcare services across sectors amplified the resilience to respond to shock; misinformation remained to be unresolved and the mutual trust of patients, professionals, and society as a whole in response to the health crisis.^27^ As the capacity of healthcare became overwhelmed, the coordination between local healthcare providers and local government was to be another challenge. To avoid recurrent outbreaks of SARS-CoV-2 after the initial pandemic wave, it was critical that the capacities of healthcare were not to exceed its saturation absent other interventions. Discussions thus far hovered over the comparatively low number of Covid-19 tests in Japan, less than 2 per 1,000 individuals compared with 12 in South Korea and 18 in the US up to the point. “Personnel-related bottlenecks” was supposed to hinder the broader use of polymerase chain reaction (PCR) to screen for the virus. In contrast, the effectiveness of pathogen testing has been confirmed in other countries.^28^ The dynamics of COVID-19 caused the intrinsic existence of individuals with undetected SARS-CoV-2 infection. It is of priority to act according to WHO’s recommendations of a combination of measures: rapid and adequate diagnosis, immediate isolation of confirmed cases, rigorous tracking and precautionary self-isolation of close contact. Medical quarantine and non-medical control have been implemented by other countries to prevent further spread and helped reduce the imported or exported cases.^18^

While it is critical to balance the control of spread and economic impact from COVID-19, when this is not feasible then priority is to be taken.^29,30^ The transmission of COVID-19 was supposed to be more infectious than past SARS outbreak.^31^ Infectiousness was estimated to peak on or before symptom onset, and thus many infections potentially took place in an unnoticeable way. Disease control measures should be adjusted to account for probable substantial subclinical transmission.^32^ COVID-19 had more severe pre-symptomatic and asymptomatic infections than influenza A and SARS and clinical studies were to evaluate the viremia and the dynamics in individuals. Heterogeneities might exist in the between-communities measures as well as in the responses to containment, growth in sporadic events could overwhelm the contact tracing system, leading to the necessity for broader-scale social interventions. Ongoing data collection, epidemiological analysis and alongside clinical research on COVID-19 are therefore essential parts of assessing the impacts of measures.^29^ Prolonged or intermittent non-medical measure such as social distancing may be of necessity years ahead.^31^ This might potentially redefine the daily routine that we are experiencing by this time.^23^

It was shown that the crude size of epidemic could be roughly estimated based on its initial dynamics under certain public health interventions. The possible trajectories of an outbreak hinged on the levels of public health interventions such as quarantine and precautionary measures.^33^ The uncertainty of the timing and duration of peak was contributable to multiple factors, including stochasticity in early dynamics, heterogeneities in contact patterns, spatial variation and dynamics of the epidemiological parameters.^29^

SARS was eventually contained by means of prompt isolation, strict quarantine of contacts, and top-down enforcement of community containment and COVID-19 outbreak of the first wave has been controlled in some countries to date. Striking similarities between SARS and COVID-19 were identified, but more difference was to be ascertained. Even if traditional public health measures are incapable to completely contain the outbreak of COVID-19, they will still be operative in reducing the peak incidence and mortality when no vaccine is available.^34^

While rigorous control policies were to associate with a slower growth in cases, in the extremity where stay-at-home restrictions are unlikely to be the one-shot deal, a gradual approach to restrictive measures might be of necessity.^23^ It will be particularly meaningful to design measures for long-term medical and non-medical control of COVID-19, along with large scale testing and contact tracing and isolation. Research should concentrate on refining specific estimates of susceptibility to infections, which is instrumental to appraise the impact of these strategies.^28^ In the absence of effective measures, pandemic spread widely and thus considerable effort at a variety of levels was of necessity for the outbreak to settle down. Such efforts will be essential to quench local outbreaks and reduce the risk of further global dissemination.^35,36^ Protective measures would compromise the effectiveness as cases accrue or the later dynamics altered significantly, in which optimization of the treatment and the development of specific medicine would be of priority when the costs of herd immunity were misestimated.^37,38^

In conclusion, the dynamics of COVID-19 incurred the intrinsic gap in the peak sizes and the peak dates between confirmed and unconfirmed cases. Interventions based on medical quarantine and non-medical containment present a strong potential to reduce the magnitude of peak sizes and cause an earlier arrival of peak dates of COVID-19 outbreak in Japan. Lowering and flattening of the pandemic peak is particularly of concern, as this reduces the acute pressure on the healthcare system as well as on the society. When it is of difficulty to pinpoint the next epidemic, the measures taken as of today would matter.^16,39^

## Data Availability

The data is in the process of uploading to GitHub.

## Contributors

NI, ZWQ, WS, and GY conceived the study. NI designed, coded the model, and made the figures. ZWQ, WS, and GY consulted on the analyses. All authors interpreted the results, contributed to writing of the article, and approved the final version for submission.

## Declaration of interests

We declare no competing interests.

## Data sharing

Data used in this study are publicly available from .the official website of Ministry of Health, Labor and Welfare of Japan.

## Acknowledgments

This research was partly funded by the FuJian Philosophy and Social Sciences Association (FJPSS)(Grant no:fj2019B08) and Science and Technology Development Center of the Ministry of Education(STDCME) (Grant no:2019J01008), using assistance from the China central and local government to support research in global health The views expressed in this publication are those of the author(s) and not necessarily those of the FJPSS or the STDCME.

## Notes

### Competing Interest Statement

The authors have declared no competing interest.

## References

1 Deslandes A, Berti V, Tandjaoui-Lambotte Y, et al. SARS-COV-2 was already spreading in France in late December 2019. J Antimicrob Agents. Available online 3 May 2020, 106006. DOI: https://doi.org/10.1016/j.ijantimicag.2020.106006

2 WHO coronavirus report. Novel coronavirus (2019-nCoV) situation report 16. World Health Organization, 2020. https://covid19.who.int.

3 Bedford J, Farrar J, Ihekweazu C, Kang G, Koopmans M, Nkengasong J. A new twenty-first century science for effective epidemic response. Nature 2019: 575, 130–136. DOI: 10.1038/s41586-019-1717-y.

4 Chen S, Yang J, Yang W, Wang C, Barnighausen T. COVID-19 control in China during mass population movements at New Year. Lancet 2020; 395: 764–66.

5 Zhu N, Zhang D, Wang W, et al. A novel coronavirus from patients with pneumonia in China, 2019. N Engl J Med 2020; published Feb 20. DOI: 10.1056/NEJMoa2001017.

6 Riou J, Althaus CL. Pattern of early human-to-human transmission of Wuhan 2019 novel coronavirus (2019-nCoV), December 2019 to January 2020. Euro Surveill 2020; 25: 2000058.

7 Prem K, Liu Y, Timothy W, et al. The effect of control strategies to reduce social mixing on outcomes of the COVID-19 epidemic in Wuhan, China: a modelling study. Lancet Public Health; published online 25 March 2020. DOI: 10.1016/S2468-2667(20)30073-6.

8 Hellewell J, Abbott S, Gimma A, et al. Feasibility of controlling COVID-19 outbreaks by isolation of cases and contacts. Lancet Glob Health 2020; 8: e488–96. Published: February 28, 2020. DOI: https://doi.org/10.1016/S2214-109X(20)30074-7.

9 Lloyd-Smith JO, Schreiber SJ, Kopp PE, Getz WM. Superspreading and the effect of individual variation on disease emergence. Nature 2005; 438: 355–59.DOI: 10.1038/nature04153.

10 Peak CM, Childs LM, Grad YH, Buckee CO. Comparing nonpharmaceutical interventions for containing emerging epidemics. Proc Natl Acad Sci USA 2017; 114: 4023-28. DOI: 10.1073/pnas.1616438114.

11 Zhao S, Lin QY, Ran JJ, et al. Preliminary estimation of the basic reproduction number of novel coronavirus (2019-nCoV) in China, from 2019 to 2020: a data-driven analysis in the early phase of the outbreak. Int J Infect Dis 2020; 92: 214–17. DOI: 10.1016/j.ijid.2020.01.050.

12 Zhang SX, Wang ZZ, Chang RJ, et al. COVID-19 containment: China provides important lessons for global response. Front Med. DOI: 10.1007/s11684-020-0766-9.

13 Wallinga J, Teunis P. Different epidemic curves for severe acute respiratory syndrome reveal similar impacts of control measures, Am J Epidemiol, 160: 6, 509–516, https://doi.org/10.1093/aje/kwh255.

14 Morens MD, Daszak P, Taubenberger KJ. Escaping Pandora’s Box — Another Novel Coronavirus. N Engl J Med 2020; 382:1293–1295.DOIa0.1056/NEJMp2002106.

15 Mizumoto K, Kagaya K, Zarebski A, Chowell G. Estimating the asymptomatic proportion of coronavirus disease 2019 (COVID-19) cases on board the Diamond Princess cruise ship, Yokohama, Japan, 2020. Euro Surveill. 2020; 25(10):pii=2000180. https://doi.org/10.2807/1560-7917.ES.2020.25.10.2000180.

16 Shigemura J, Ursano RJ, Morganstein JC, Kurosawa M, Benedek DM. Public responses to the novel 2019 coronavirus (2019-nCoV) in Japan: Mental health consequences and target populations. Psychiatry Clin Neurosci. 74: 4, 281–282.DOI: 10.1111/pcn.12988.

17 Wu JT, Leung K, Leung GM. Nowcasting and forecasting the potential domestic and international spread of the 2019-nCoV outbreak originating in Wuhan, China: A modelling study. Lancet 2020. https://doi.org/10.1016/S0140-6736(20)30260-9.

18 Anzai A, Kobayashi T, Linton, NM, et al. Assessing the Impact of Reduced Travel on Exportation Dynamics of Novel Coronavirus Infection (COVID-19). J Clin Med.9: 2, 601.DOI: 10.3390/jcm9020601.

19 Ministry of Health, Labor and welfare. Declaration of a state of emergency in response to the novel coronavirus disease. https://www.mhlw.go.jp/content/10900000/000624434.pdf.

20 Maier BF, Brockmann D. Effective containment explains subexponential growth in recent confirmed COVID-19 cases in China. Science. 2020 Apr 8. pii: eabb4557. DOI: 10.1126/science.abb4557.

21 Anastassopoulou C, Russo L, Tsakris A, Siettos C. Data-based analysis, modelling and forecasting of the COVID-19 outbreak. PLoS One. 2020 Mar 31; 15(3):e0230405. DOI: 10.1371/journal.pone.0230405.

22 Rong XM, Yang L, Chu HD, Fan M. Effect of delay in diagnosis on transmission of COVID-19. Math Biosci Eng. 2020 Mar 11; 17(3):2725–2740.DOI: 10.3934/mbe.2020149.

23 Studdert DM, Hall MA. Disease Control, Civil Liberties, and Mass Testing—Calibrating Restrictions during the Covid-19 Pandemic. N Engl J Med. April 9, 2020.DOI: 10.1056/NEJMp2007637.

24 Ferguson NM, Keeling MJ, Edmunds WJ, et al. Planning for smallpox outbreaks. Nature 425, 681–685 (2003). DOI: https://doi.org/10.1038/nature02007.

25 Sjodin H. Wilder-Smith A, Osman S, Farooq Z, Rocklöv J. Only strict quarantine measures can curb the coronavirus disease (COVID-19) outbreak in Italy, 2020. Euro Surveill. 2020 Apr; 25(13). DOI: 10.2807/1560-7917.ES.2020.25.13.2000280.

26 Svoboda T, Henry B, Shulman L, et al. Public health measures to control the spread of the severe acute respiratory syndrome during the outbreak in Toronto. N Engl J Med. 2004 Jun 3; 350(23):2352–61. DOI: 10.1056/NEJMoa032111.

27 Legido-Quigley H, Asgari N, Teo YY, et al. Are high-performing health systems resilient against the COVID-19 epidemic? Lancet. 2020 Mar 14; 395(10227):848–850. doi: 10.1016/S0140-6736(20)30551-1.

28 Zhang J, Litvinova M, Liang Y, et al. Changes in contact patterns shape the dynamics of the COVID-19 outbreak in China. Science. 29 Apr 2020. DOI: 10.1126/science.abb8001.

29 Anderson RM, Heesterbeek H, Klinkenberg D, Hollingsworth TD. How will country-based mitigation measures influence the course of the COVID-19 epidemic? Lancet. 2020. 2020 Mar 21; 395 (10228):931–934. DOI:http://dx.doi.org/10.1016/S0140-6736(20)30567-5.

30 Cobey S, Modeling infectious disease dynamic. Science 24 Apr 2020:eabb5659. DOI: 10.1126/science.abb5659.

31 Kissler SM, Tedijanto C, Goldstein E, Grad YH, Lipsitch M. Projecting the transmission dynamics of SARS-CoV-2 through the postpandemic period. Science 2020:eabb5793.DOI: 10.1126/science.abb5793.

32 He X, Lau EHY, Wu P, et al. Temporal dynamics in viral shedding and transmissibility of COVID-19. Nat Med. 2020 Apr 15. doi: 10.1038/41591-020-0869-5.

33 Nishiura H, Patanarapelert K, Sriprom M, Sarakorn W, Sriyab S, Ming Tang I. Modelling potential responses to severe acute respiratory syndrome in Japan: the role of initial attack size, precaution, and quarantine. J Epidemiol Community Health. 2004 Mar; 58(3):186–91.DOI: 10.1136/jech.2003.014894.

34 Wilder-Smith A, Chiew CJ, Lee VJ. Can we contain the COVID-19 outbreak with the same measures as for SARS? Lancet Infect Dis. 2020 Mar 5. pii: S1473-3099(20)30129-8. https://doi.org/10.1016/S1473-3099(20)30129-8.

35 Lipsitch M, Cohen T, Cooper B, et al. Transmission Dynamics and Control of Severe Acute Respiratory Syndrome. Science 2003; Jun 20; 300(5627):1966–70. DOI: 10.1126/science.1086616.

36 Verity R, Okell CL, Dorigatti I, et al. Estimates of the severity of coronavirus disease 2019: a model-based analysis. Lancet Infect Dis. 2020 Mar 30. pii: S1473-3099(20)30243-7. DOI: https://doi.org/10.1016/S1473-3099(20)30243-7.

37 Fang Y, Nie Y, Penny M. Transmission dynamics of the COVID-19 outbreak and effectiveness of government interventions: A data-driven analysis. J Med Virol. 2020 Mar 6. DOI: 10.1002/jmv.25750.

38 Li Q, Guan X, Wu P, et al. Early Transmission Dynamics in Wuhan, China, of Novel Coronavirus–Infected Pneumonia. N Engl J Med 2020; published online Jan 29 382:1199–1207. DOI: 10.1056/NEJMoa2001316.

39 Gurdasani D, Ziauddeen H. On the fallibility of simulation models in informing pandemic responses. Lancet Glob Health. Published: April 30, 2020. DOI: https://doi.org/10.1016/S2214-109X(20)30219-9.

